# A Passive-State Comparison of Shear Wave Ultrasound Elastography of the Biceps Brachii with a Robotic Measurement of Elbow Extension Impedance in Chronic Hemiparetic Stroke

**DOI:** 10.1101/2020.07.15.20154658

**Authors:** Michael D. Ellis, Ninette T. A. Gerritsen, Netta Gurari, Sabrina M. Lee, Julius P. A. Dewald

## Abstract

Muscle tissue is prone to changes in composition and architecture following stroke. Changes in muscle tissue of the extremities are thought to increase passive muscle stiffness and joint impedance. These effects likely compound neuromuscular impairments exacerbating movement function. Unfortunately, conventional rehabilitation is devoid of quantitative measures yielding to subjective assessment of passive joint mobility and end feel. Shear wave ultrasound elastography is a conventional tool used by ultrasonographers that may be readily available for use in the rehabilitation setting as a quantitative measure, albeit at the muscle-tissue level, filling the gap. To support this postulation, we evaluated the *criterion* validity of shear wave ultrasound elastography of the biceps brachii by investigating the relationship to a laboratory-based criterion measure for quantifying elbow joint impedance in individuals with moderate to severe chronic stroke. Measurements were performed under passive conditions at seven positions spanning the arc of elbow joint extension in both arms of twelve individuals with hemiparetic stroke. Surface electromyography was utilized for threshold-based confirmation of muscle quiescence. A significant moderate relationship was identified near end range of elbow extension and all metrics were greater in the paretic arm. Data supports the progression toward clinical application of shear wave ultrasound elastography in evaluating altered muscle mechanical properties in stroke stipulating the confirmation of muscle quiescence. Considering the lack of bedside robotics in clinical practice, shear wave ultrasound elastography will likely augment the conventional method of manually testing joint mobility. Tissue-level measurement may also assist in identifying new therapeutic targets for patient-specific impairment-based interventions.

**New & Noteworthy:** Methods for quantifying passive (non-reflex mediated) joint mobility are absent in stroke rehabilitation. Rehabilitation specialists are left to subjective assessment of the impact on function. Here, we compare the application of shear wave ultrasound elastography for estimating mechanical properties of muscle with a robotic method (criterion measure) of measuring passive elbow extension joint impedance. Data support the clinical application of shear wave ultrasound elastography, especially with the absence of bedside robotics.

## Introduction

Methods for the quantitative evaluation of passive (not active/reflexive, eg. spasticity) resistance to passive range of motion are absent in the stroke rehabilitation setting leaving clinicians to subjective assessment. This is problematic, especially in the later stage of stroke recovery that is notable for a loss of independent joint control (17) which devastates reaching range of motion (73). The diminished activity and use of the arm (23) that accompanies the loss of independent joint control places patients at risk for secondary musculoskeletal changes such as muscle atrophy (24), reduced fascicle length (42), lengthened sarcomeres (50), and accumulation of connective tissue or muscle fibrosis (45). These muscle tissue changes threaten to compound neuromuscular impairments of reaching range of motion, like loss of independent joint control that results in decreased elbow extension excursion and velocity due to the resistance imparted by abnormal flexor activation (22). In fact, the passive mechanical contributions of resistance to joint rotation are increased at the ankle (16, 28, 30, 47, 49, 54, 56) and elbow (4, 30, 51, 54, 56) in individuals with stroke. More specifically and relevant to reaching function, passive elbow extension joint impedance increases towards the end of the range of motion (4, 14, 49, 54, 55). Importantly, the joint-level impairment reflects the tissue-level exponential passive stress-strain relationship that is reported for muscle fascicles (1, 62, 64), highlighting the role of muscle tissue stiffness in passive joint impedance. Unfortunately, methods for the measurement of passive end-range joint impedance that would benefit clinical decision making have been restricted to laboratory-based investigations, limiting clinicians to subjective manual evaluation of joint mobility and end-feel (61). Objective tools for clinical evaluation are needed in order to improve rehabilitation diagnoses of physiologic mechanisms underlying reaching dysfunction following stroke.

Clinical evaluation at the tissue level is an alternative approach and may provide more precise mechanistic information and, therefore, identify potential therapeutic targets for the development of impairment-based rehabilitation interventions. The measurement of passive joint impedance represents the net effect of all tissues surrounding the joint including, but not limited to muscles, ligaments, and joint capsules (65); however, muscle tissue is thought to be the major contributor (44). Integrating interventions targeting stroke-related muscle tissue changes may augment current impairment-based interventions that are aligned with restoring premorbid neural control of movement. For example, the prominent expression of a flexion synergy and impaired reaching range of motion in severe stroke (23) suggests that elbow flexors may be the primary contributor to joint-level changes in passive impedance. Therefore, targeting stroke-related changes in biceps brachii may augment interventions for the flexion synergy, such as progressive abduction loading therapy (20). Successful administration of this approach would require evaluation at the tissue level. However, until recently, the only technique available to quantify the mechanical properties of individual muscles is magnetic resonance elastography (46), which is too costly to justify in stroke rehabilitation.

A prime candidate for measurement of mechanical properties at the muscle tissue level in the clinical setting is shear wave ultrasound elastography. Shear wave ultrasound elastography is a promising solution to enhancing the diagnostic capabilities of stroke physicians and rehabilitation specialists since it is routinely utilized in conventional practice for imaging other tissues such as breast (7) and liver (26). Shear wave ultrasound elastography can be employed using a conventional diagnostic ultrasound device equipped with shear wave capabilities to measure shear wave velocity in muscle (34). Shear wave velocity is proportional to the square root of the shear modulus and, therefore, is often reported as a surrogate measure for the elastic properties of the tissue (8, 13, 19), i.e. “muscle stiffness”. In brief, the technology uses a piezoelectric array to successively focus the depth of the acoustic beam such that the focus travels into the muscle at supersonic speeds (8). The supersonic speed of the acoustic beam focus results in shear waves propagating in opposite orthogonal directions from a Mach cone at a much slower speed (1-2m/s) than the shear source (Mach 3-4). High frame-rate imaging (3kHz-6kHz) interspersed between shear source/push beam sequences allow for the measurement of the shear wave propagation velocity.

Evidence for *face* validity of shear wave velocity as a surrogate measure of muscle stiffness in passive muscle has been demonstrated in a cadaveric animal preparation employing a direct stress/strain-based calculation of the Young’s modulus (19). An *in situ* feline study has demonstrated that under active conditions, shear wave velocity is sensitive to changes in net material properties of muscle (Bernabei et al., 2020). Shear wave ultrasound elastography has been routinely employed to investigate neurologically intact muscle under both passive and active conditions (10-12, 60). Furthermore, ultrasound elastography has been used to evaluate the effects of hemiparesis, demonstrating increased muscle stiffness in both cerebral palsy (33) and stroke (38, 39) supporting the *face* validity. Evidence for the *criterion* validity of ultrasound elastography exists for neurologically intact humans but in the ankle joint using robotic-based measurement of passive joint impedance as a criterion measure (14). Evidence is limited however supporting the *criterion* validity of ultrasound elastography in hemiparetic biceps with only one study to-date (18) comparing shear wave velocity measurements to a criterion measure of joint impedance. While the exploratory study by Eby et. al. (18) supports the *criterion* validity of ultrasound elastography, the results are limited in that half of the sample (n=4 of 9) were mildly impaired such that they presented similarly to controls. These data warrant further investigation of a larger sample, especially of individuals with homogeneous moderate/severe impairment.

Therefore, the primary aim of this study was to evaluate the *criterion* validity of ultrasound elastography for assessing muscle stiffness following stroke. The relationship between biceps ultrasound elastography metrics (shear wave velocity, shear modulus) and elbow extension joint impedance was investigated using a custom one-degree-of-freedom device (25) at seven angular positions. Joint impedance was chosen as the criterion measure, similar to Eby et. al. (18) as well as in related measurements such as myotonometry (43). Joint impedance is considered in lieu of alternative criterion measures, such as magnetic resonance elastography, since it represents a quantitative analogue to what is manually performed by the clinician when evaluating passive joint mobility. In the present study, we hypothesized that there would be a positive/direct relationship between the two measurement techniques since the biceps brachii substantially contributes to the net joint torque exerted about the elbow joint. A stronger correlation between the metrics was expected at the end range of motion due to the simultaneous lengthening of flexors and shortening of extensors that would respectively increase and decrease their contribution to the net torque exerted about the elbow joint. The results of this investigation provide evidence for the *criterion* validity of ultrasound elastography in hemiparetic muscle and support its application in clinical practice.

## Materials and Methods

### Participants

A total of twelve individuals (six men, six women) age 57±10 years with chronic stroke (13±9 years post) and upper extremity Fugl-Meyer Motor Assessment scores of 27±3 out of 66, participated in and completed the study. All participants were recruited from the Clinical Neuroscience Research Registry of Shirley Ryan AbilityLab and Department of Physical Therapy and Human Movement Sciences of Northwestern University. The inclusion criteria for individuals with chronic stroke were: (1) hemiparetic stroke with a moderate to severe impairment of the upper limb as determined by clinical evaluation using the Arm Motor Fugl-Meyer Assessment, (2) adequate passive range of motion at the shoulder and elbow joints without pain at end range, (3) ability to provide informed consent.

Prior to the experiment, all participants gave informed consent to participate in the study. The study was approved by the Institutional Review Board of Northwestern University in accordance with the ethical standards laid down in the 1964 Declaration of Helsinki for research involving human subjects.

Participants who were included were first screened by a licensed physical therapist (Ellis) to confirm pain-free passive range of motion at the shoulder and elbow. The shoulder was required to have passive abduction (frontal plane elevation) to at least 100°, and the elbow was required to have passive extension to at least 30° less than full normal range without pain at end range with over-pressure. No participants were excluded for conditions such as: (1) pain at end range with over-pressure indicating inflammation or (2) current pain in the extremities or spine.

### Experimental setup

The participant was positioned in the Biodex chair (Biodex Medical Systems, Inc., Shirley, New York, USA), with the arm/shoulder abducted 90°, arm/shoulder rotated 45° from the coronal plane, and the elbow at 90° (Figure 1). The participant was strapped into the Biodex chair with nylon belts to constrain movement of the upper body and shoulder girdle. The forearm, including wrist and fingers, were casted with fiberglass and indirectly coupled to a one-dimensional tension-compression load sensor (Stamford, CT, USA, Omega Engineering Inc., LCM201-300) via a custom one-degree-of-freedom robotic device (25).

**Figure 1.**
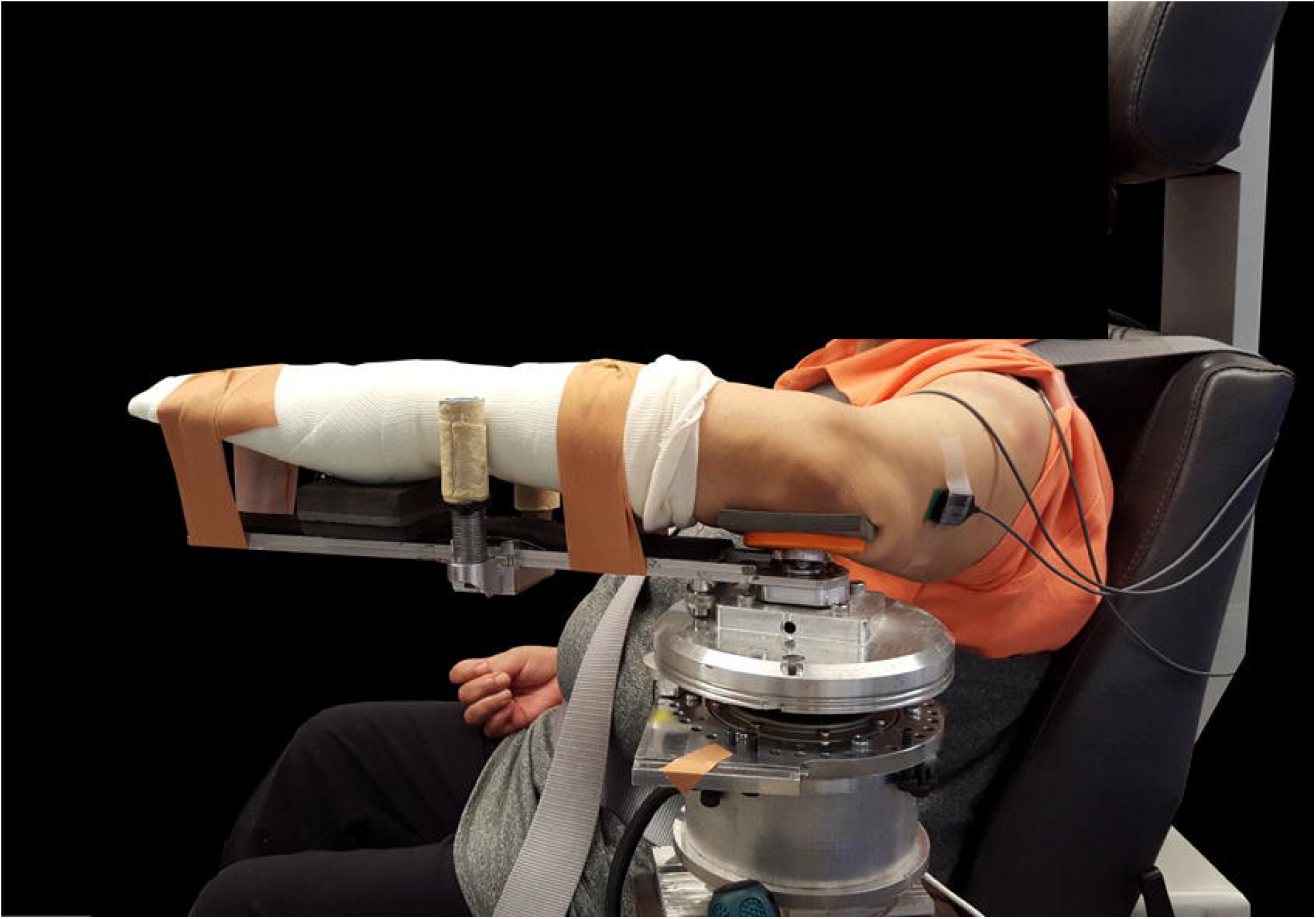
Experimental setup. The lower arm is casted and mounted comfortably to the swing arm of the robotic device in the standard arm configuration. The torso is secured to the seat with nylon strapping. EMG electrodes are in place on the biceps brachii and the long head of the triceps brachii.

The fiberglass casting allowed for a non-invasive rigid coupling of the participant’s forearm to the device, encouraging direct force transmission from the participant’s forearm to the load sensor. Casting also constrained the pronation/supination degree-of-freedom. The medial epicondyle was physically palpated and placed in the middle of a padded cup, such that the axis-of-rotation of the elbow was aligned with the axis-of-rotation of the robotic device. The robotic device controlled the elbow joint angular velocity and concurrently measured the torque about the elbow joint. An encoder (Peabody, MA, USA, US250), that was attached to the device’s motor, measured the angular position of the elbow joint. Shear wave velocity images were captured using an ultrasonography system (Aixplorer SuperSonic Imagine, Aix en Provence, France) with a linear transducer array (4–15MHz, SuperLinear 15–4, Vermon, France). Surface EMG (Delsys, Bagnoli EMG System, Boston, MA, USA) was recorded throughout the session for the biceps brachii and the long head of the triceps brachii. Torque/angle data were sampled at 1kHz and stored offline for post hoc analysis. Surface EMG data were collected with a Delsys 4-channel Bagnoli EMG system (Boston, MA). Active differential electrodes with a 1cm inter-electrode distance were used. EMG signals were pre-filtered with a low-pass cutoff frequency of 500Hz (8-pole Butterworth filter) and amplified by 1000.

### Protocol

The participant was first positioned in the standardized configuration as described in the experimental setup section. Once positioned, the elbow joint was manually flexed and extended by the experimenter over the entire range of motion to confirm proper alignment and comfortable full range of motion at the elbow joint upon a stationary humerus. EMG sensors were placed on prepped skin over the medial muscle belly of biceps brachii and the long head of the triceps brachii. The ground sensor was placed proximally at the acromioclavicular joint. The participant’s elbow joint was then placed in a standardized position of 90°, and the software recorded this position as the 90° home position.

A standardized reflex habituation procedure was then administered to minimize muscle stretch reflexes across participants. In this procedure, the elbow joint was passively rotated between 75° and 150° (elbow angle) at 120°/s for twenty repetitions. The angular velocity was sufficient to elicit repetitive stretch reflex activity resulting in a habituation of abnormal tonic muscle activation (69). If muscle activation was present after the stretches, visualized as spikes in the steady-state EMG signal while at rest, stretches were repeated. If quiescence was confirmed, a subsequent 5s recording was made and stored offline for future analysis to define the “baseline” quiescent EMG signal. Two participants were unable to achieve quiescence in their muscle activity leading to termination of the experiment and reduction of the study sample to N=10. After the habituation procedure, the torque signal was zeroed with the participant at rest in the standardized 90° elbow joint position.

A standardized procedure was then administered for the acquisition of passive elbow joint torque during continuous elbow joint rotation by the device. In this procedure, the device rotated the elbow at a constant angular velocity of 6°/s from 75° to 150° (elbow angle), an angular velocity slow enough as to not elicit a hyperactive stretch reflex (30). The 75° boundary was the most acute elbow angle that could be safely implemented in the robotic setup while the 150° boundary was implemented empirically to avoid length-related reflex muscle activation that would violate the passive/quiescent requirements of the study. In one participant, the final elbow angle was reduced to 140° due to minor discomfort at the 150° position despite having a greater pain-free passive range of motion during initial screening. A 1s hold occurred at end-range of extension prior to returning the segment back to the starting position. A 1s hold occurred prior to the onset of the subsequent trial with ten repetitions being completed in approximately 4.5 minutes.

The next step of the protocol involved capturing ultrasound elastography images of the biceps brachii at seven different elbow joint angle positions (Figure 2). The most acute elbow angle measured was 90° and proceeded with 100°, 110°, 120°, 130°, 140° and 150°. The 90° position was the most acute angle that could be measured due to the size of the ultrasound transducer and its proximity to adjacent anatomy. At each angular position, five images were collected that passed the following criteria: 1) muscle fascicles were aligned in the plane of the transducer, 2) the shear wave velocity map region of interest (blue box in Figure 2) was placed in the mid-belly region distal and superficial to the central aponeurosis of the biceps, 3) the shear wave velocity map region of interest illustrated acquisition of velocity data throughout the entire area, 4) no muscle activity was visually observed from either of the muscles, 5) minimal transducer pressure was applied to the muscle, and 6) images were captured 45s after placing the joint in the subsequent angular position (32). Visual inspection of the electromyography signal for muscle activation and participant posture was performed throughout the experiment. An output signal from the ultrasound system indicating image capture was recorded with the torque, position, and EMG data; the EMG signals were analyzed offline to confirm muscle quiescence. The protocol originally intended to also measure shear wave velocity in the lateral head of the triceps. However, imaging of the triceps was dropped from the protocol following the inability to meet criteria #3 above in the first five participants.

**Figure 2.**
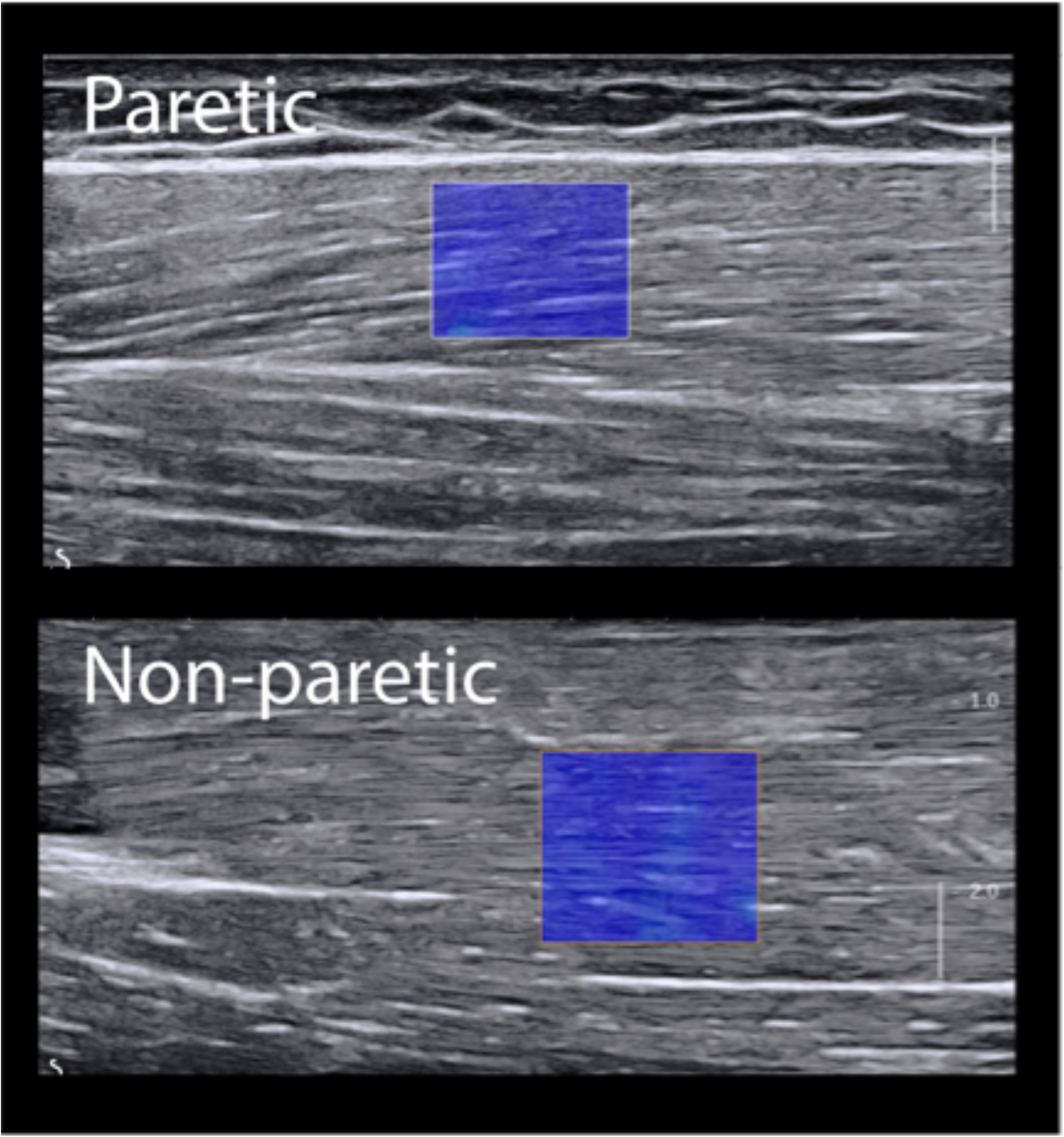
Shear wave ultrasound elastography images for the paretic and non-paretic side at the same joint angle (140°). The paretic biceps muscle has a mean ± standard deviation shear wave velocity of 4.55 ± 0.033m/s. The non-paretic image has a shear wave velocity of 2.824 ± 0.01m/s. The blue box represents the region of interest.

The final step in the protocol was to determine maximum muscle activation. The participant’s forearm was taken out of the robotic device. The participant remained strapped in the Biodex chair, and their tested arm was manually held in the standardized configuration. The maximal activity of the biceps and triceps brachii muscles was measured by requesting the participant to generate their maximal isometric elbow flexion and extension torque while the experimenter maintained the participant in the standardized configuration. Maximal voluntary torque generation in each direction was repeated twice. Diligent visual inspection of muscle activation and participant posture was maintained throughout the experiment.

### Data Analysis

EMG signals were pre-filtered with a low pass cutoff frequency of 500Hz (8-pole Butterworth filter) and amplified by 1000. Processing of EMG signals consisted of DC offset correction and full-wave rectification. Then the processed EMG signals were filtered using a zero-phase 250ms moving average window. Quiescent muscle activation was defined as the filtered EMG signal remaining below the threshold of mean plus three times the standard deviation (66). The threshold was calculated from the “baseline” EMG data acquisition trials recorded for each muscle in each experimental session (paretic or non-paretic arm). The threshold for each muscle was then applied across the entire data set of the experimental session to confirm muscle quiescence during acquisition of both passive joint torque and shear wave velocity.

Processing of the torque data began with filtering using a zero-phase 250ms moving average window. Position-based averaging of the ten repetitions of slow stretches was then conducted for each participant resulting in a mean torque-angle curve. Next, the torque data were cropped at the beginning and end of the trial to retain data for only the constant velocity portion of the trial and to remove angular acceleration artifacts (30). This resulted in the analysis of the angular excursion from 77.5° to 147.5°. Post-processing proceeded with fitting a 3^rd^ order polynomial model, *τ*(*θ*) = *p*_1_*θ*^3^ + *p*_2_*θ*^2^ + *p*_3_*θ* + *p*_4_, to the position-averaged data. The fit was accepted if the variance accounted for, as determined by R^2^, was above 0.98. This approach reflected prior methods (70, 72), and most importantly, allowed joint impedance to be calculated at the seven specific joint positions where ultrasound measurements were taken. Furthermore, this approach allowed joint impedance to be calculated at more acute and obtuse joint angles as opposed to evaluating only the mid-range linear portion as conducted in prior work (28, 30, 43) or by fitting the torque response to a second order mass-spring-damper model (51) where stiffness is assumed constant. Model parameters (*p*_1_, *p*_2_, *p*_3_, *p*_4_) were determined with the function ‘fit’ provided by MATLAB (MathWorks, Inc). Joint impedance was determined by taking the analytical derivative of the function (*τ*′(*θ*) = 3*p*_1_*θ*^2^ + 2*p*_2_*θ* + *p*_3_) and inputting the model parameters. Specifically, the joint impedance was calculated for each angular position by implementing *θ* = 90°, 100°, 110°, 120°, 130°, 140°, and 150° in the derivative function equation. Since the model is fit on data that only contained a robot-commanded constant velocity, the 150° position was extrapolated from the model (Figure 3).

**Figure 3.**
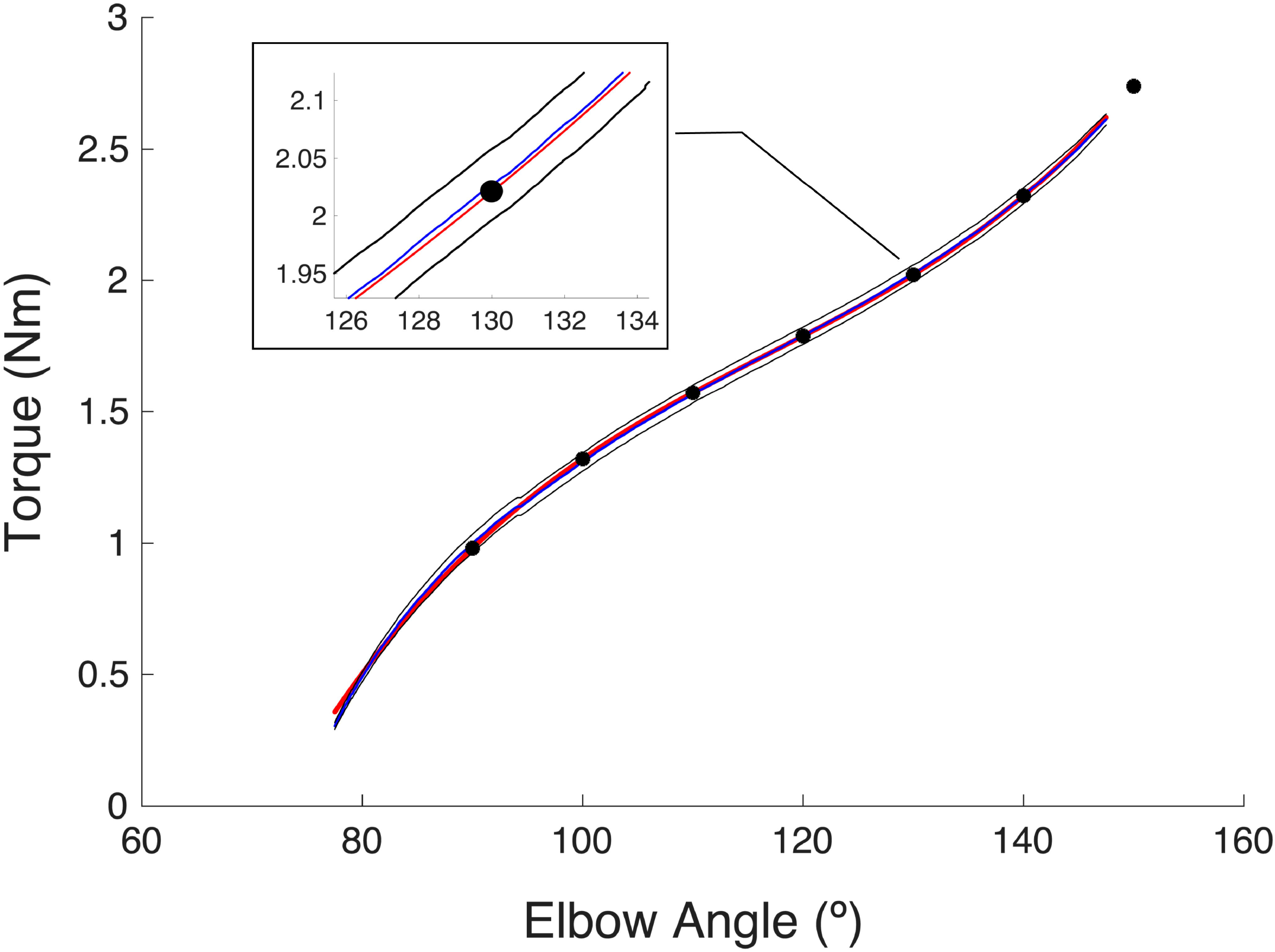
Position-averaged torque profile (blue line), 2 standard deviation cloud (black lines), and 3^rd^ order polynomial fit (red line). The 7 tested joint angles are indicated with black dots. A 2.5° gap is visible before 150° due to the data exclusion of the acceleration profile.

As mentioned above, muscle quiescence was confirmed throughout data acquisition for the joint impedance protocol. Specifically, within each repetition, a range of 4° (*θ*-2° to *θ*+2°) was taken at each standardized angular position and analyzed to determine if both filtered EMG signals remained below their respective threshold. If muscle activation was identified, the repetition was excluded from the average since it would invalidate the passive measurement. This occurred in 3 out of the 120 total trials in the protocol.

Post-processing of the ultrasound data was performed with custom software written in MATLAB (Mathworks, Natick, USA) (39). The custom software extracts the shear wave velocity and quality factor, producing two 60 × 60 matrices for the region of interest (ROI, i.e. blue box in Figure 2). The company software provides a proprietary ‘quality factor’ value to indicate the ‘quality’ of each shear wave velocity measurement. This is related to the cross-correlation algorithm that the company software uses to track the propagation of shear waves through the tissue. Mean shear wave velocity was then calculated from the velocity values within the ROI that surpassed the device-default 0.7 quality factor for each of the five trials within each of the seven joint positions. Shear wave velocity was converted to shear modulus in order to report the relationship of both shear wave velocity and shear modulus to joint impedance, allowing for better comparison with other published work (14, 39). The formula used for calculating shear modulus was, *μ* = *ρc*^2^, where *μ* is the shear modulus (kPa), *ρ* is the density of muscle, and *c* is the shear wave velocity (8).

*F*inally, the EMG signals that corresponded with each ultrasound image were analyzed for the 200ms window prior to capturing the image to confirm the absence of muscle activity that would impact shear wave velocity. Images were discarded if either of the two filtered EMG signals were above threshold as described above. An example of EMG data corresponding to ultrasound image capture is illustrated in Figure 4. Occasionally, there was a signal artifact from the ultrasound transducer (1Hz) that occurred when the ultrasound transducer was close to the EMG sensor. EMG data were visually inspected to ensure that trials were not rejected due to the presence of the artifact during the 200ms window. Instead, a broader window of 2s was used for visualization to gain insight from the preceding and proceeding EMG signals surrounding the image capture window. The image was discarded only if it was determined that the source of the increased EMG signal was from muscle activation and not the transducer artifact. Of the entire data set, only 8 of ∼700 images were rejected due to muscle activation.

**Figure 4.**
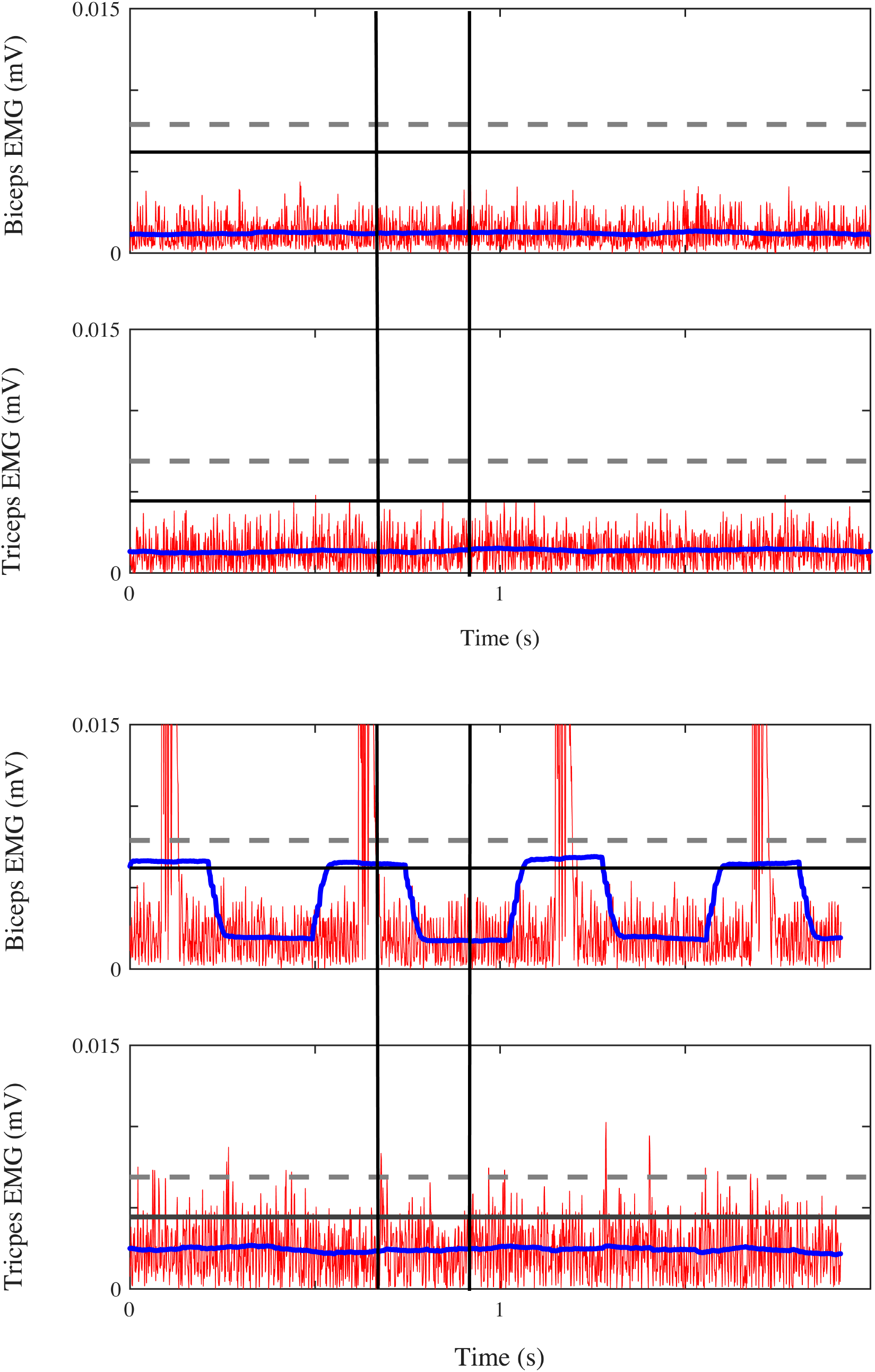
Example EMG data illustrating the post-processing threshold-based confirmation of muscle quiescence. EMG signals corresponding to a 2s time frame of interest are displayed. Two black vertical lines depict 200ms surrounding the ultrasound image capture. Significant muscle activation is not detected in either example for biceps or triceps despite the presence of the acoustic ultrasound artifact. Processed EMG data (DC-offset correction and full-wave rectification) are indicated in red, processed EMG data (zero-phase 250ms moving window filter) in blue, and EMG quiescence threshold in black (horizontal line). For greater perspective of the EMG magnitude, a threshold representing 3% maximum voluntary activation is indicated with a horizontal dotted line (grey).

### Statistical analysis

Dependent variables of mean shear wave velocity, shear modulus, and joint impedance for each of the seven joint positions were analyzed with IBM SPSS Statistics Version 24 with an alpha-level of 0.05. Prior to the analysis, a normal distribution was identified for all variables using quantile-quantile plots. Sphericity was assessed using Maukley’s Test prior to all analyses of variance.

A Pearson correlation coefficient was used to assess the relationship with joint impedance for both shear wave velocity and shear modulus. First, a global correlation assessment was performed combining all elbow positions (90°, 100°, 110°, 120°, 130°, 140°, and 150°), producing a sample of n=138 (10 participants × 2 arms × 7 positions; 2 participants were removed from the 90° condition due to body size with inadequate space for the ultrasound probe). In order to test if the correlation between joint impedance and shear wave velocity and shear modulus were increased towards the end range in elbow extension, Pearson correlation tests were conducted for subsets of angle ranges. The subsets included: 100° to 150° (n=120), 110° to 150° (n=100), 120° to 150° (n=80), 130° to 150° (n=60), 140° to 150° (n=40), and 150° (n=20).

Three separate two-way repeated measures analysis of variance (ANOVA) tests were used to test the main effects of arm (paretic/non-paretic) and joint angle (90°, 100°, 110°, 120°, 130°, 140°, and 150°) on shear wave velocity, shear modulus, and joint impedance.

## Results

There was a significant mild correlation of shear wave velocity (*r* = 0.37, n = 138, p < 0.01), and shear modulus (*r* = 0.37, n = 138, p < 0.01) with joint impedance for the combined 90°-150° data set. Figure 5 illustrates a scatter plot of shear wave velocity as a function of joint impedance for the entire data set. Subsequent stratified correlation tests listed in Table 1 indicated significant increasing *r*-values with a positive moderate correlation near end range of motion for the 150° data set (*r* = 0.49, n = 20, p < 0.01).

**Figure 5.**
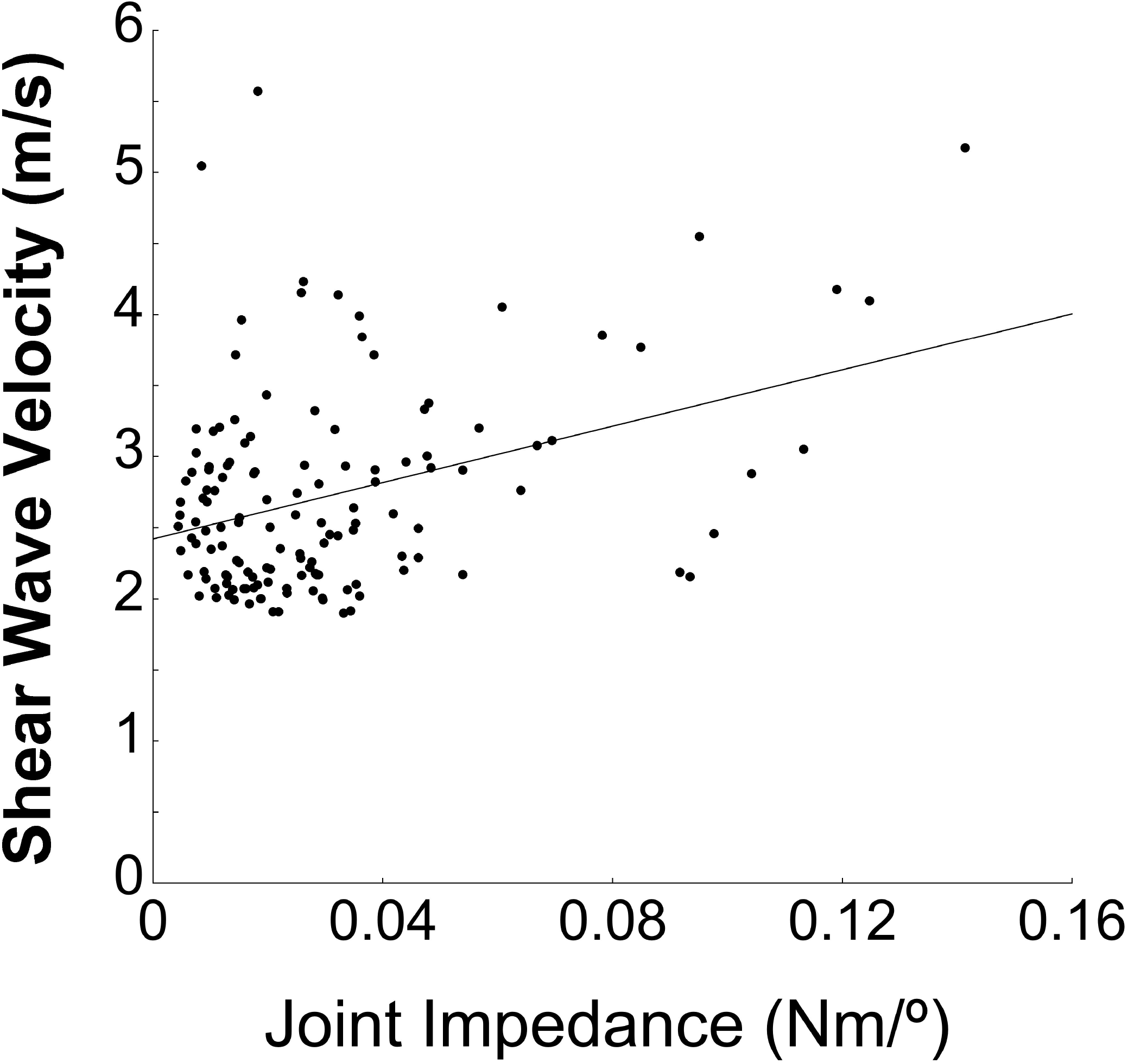
Shear wave velocity by joint impedance scatter plot for the entire data set, illustrating the data spread. A best-fit line is indicated for visual appreciation only.

**Table 1.** Statistical summary table of Pearson correlation coefficients (*r*) for the evaluation of joint impedance with shear wave velocity and shear modulus for each progressive data set.

| **Data Set** | ***Shear Wave***<br>***Velocity*** | ***Shear***<br>***Modulus*** | **n** | **p** |
| --- | --- | --- | --- | --- |
| 100° to 150° | 0.397 | 0.391 | 120 | <0.01 |
| 110° to 150° | 0.426 | 0.412 | 100 | <0.01 |
| 120° to 150° | 0.453 | 0.430 | 80 | <0.01 |
| 130° to 150° | 0.451 | 0.425 | 60 | <0.01 |
| 140° to 150° | 0.452 | 0.422 | 40 | <0.01 |
| 150° | 0.493 | 0.491 | 20 | <0.01 |

There was a significant effect of arm for biceps shear wave velocity (p = 0.05), shear modulus (p = 0.05), and joint impedance (p = 0.02) (Figure 6). The mean and standard deviation of joint impedance across all conditions was 0.04±0.03Nm/° on the paretic side and 0.02±0.01Nm/° on the non-paretic side. The biceps brachii exhibited a mean shear wave velocity on the paretic side of 2.81±0.56m/s and on the non-paretic side of 2.15±0.15m/s. The biceps brachii exhibited a mean shear modulus on the paretic side of 8.47±3.62kPa and on the non-paretic side of 5.44±0.76kPa. There was also a significant effect of joint angle for shear wave velocity (p < 0.01), shear modulus (p < 0.01), and joint impedance (p = 0.02).

**Figure 6.**
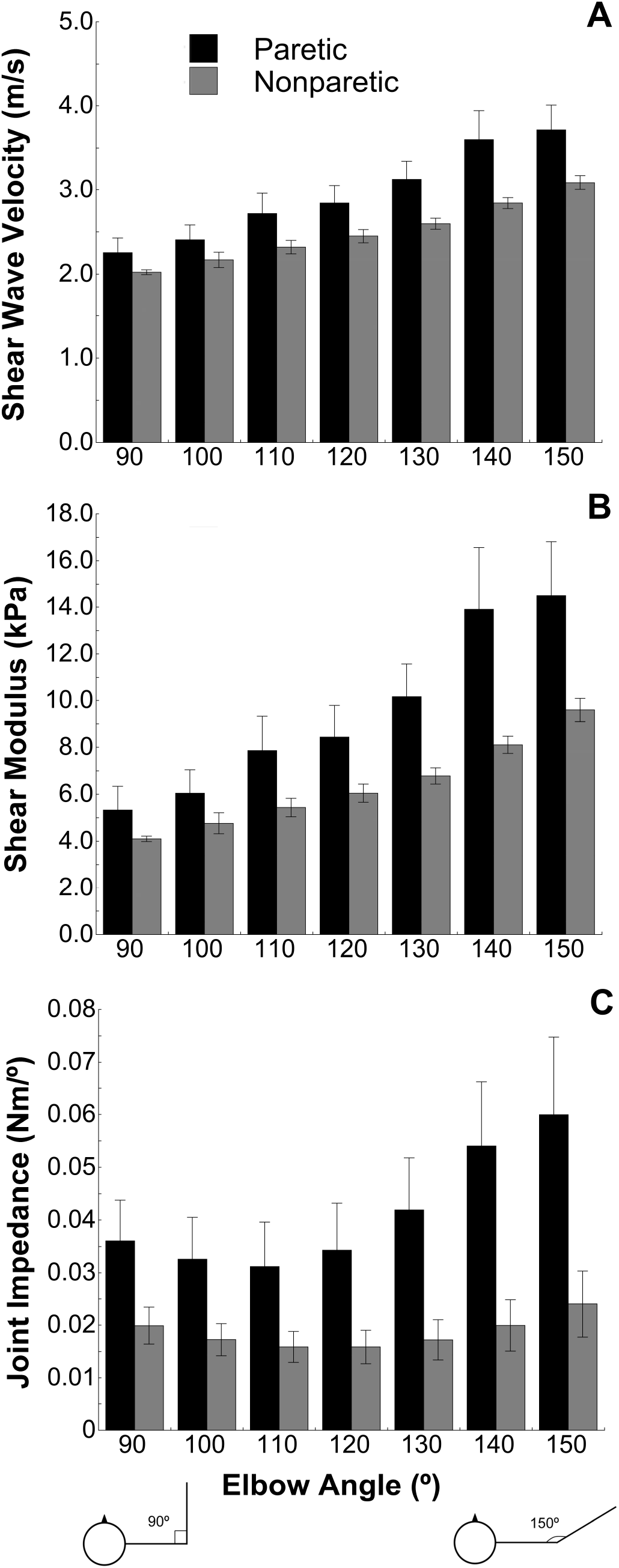
Summary data for shear wave velocity, shear modulus, and joint impedance for each tested elbow angle. The stick figure is presented at the bottom for clarification indicating the progression from acute to obtuse elbow angles reading when reading from left to right. There was a significant effect of arm and elbow angle for all three variables.

## Discussion

The moderate correlation (*r* = 0.49) found at 150° for joint impedance and muscle-tissue metrics supports the *criterion* validity of ultrasound elastography for assessing muscle-level contributions to joint mobility following stroke. It is important to acknowledge that the criterion measure in our analysis, joint impedance, has inherent differences that engender prudence in our interpretation of the moderate *r*-value as evidence for the *criterion* validity of ultrasound elastography. Joint impedance is a global measure of resistance to passive angular joint motion whereas ultrasound elastography is related to the muscle mechanical properties. The primary assumption justifying the selection of this criterion measure is that muscle tissue is assumed to be the greatest contributor to joint-level resistance to passive motion (44). Furthermore, and key to the objective of this study, is that the robotic measurement of joint impendence is a quantitative analogue to manual assessment of joint mobility and end feel, the conventional clinical assessment we hope to improve upon. Our hypothesis was supported in that the relationship between joint impedance and muscle stiffness metrics strengthened at more obtuse elbow angles where the primary contributor to passive resistance is tensioned elbow flexors. A detailed discussion of joint impedance, shear wave velocity, and shear modulus as a function of elbow angular position follows.

Joint impedance, shear wave velocity, and shear modulus demonstrated differing relationships with elbow joint angular position perhaps explaining the mild relationship for the global data set (90° to 150°) in comparison to the moderate relationship near end range of motion (150°). Joint impedance demonstrated a non-linear or parabolic relationship reflecting prior work in individuals with stroke (54). In contrast to joint impedance, shear wave velocity and shear modulus of the biceps appeared monotonic and linear as a function of joint angle. The parabolic shape of the joint impedance profile as a function of elbow angle may be explained by the antagonistic contributions of the flexor and extensor muscles as a function of elbow angle. Consider the contributions of flexor and extensor muscles to net joint torque that is used to calculate impedance; in a flexed elbow position, the triceps brachii is in a lengthened position while the biceps brachii is in a shortened position acute to its slack angle. The slack angle of biceps brachii, defined as the angle at which passive stiffness increases, is approximately 95° (35). Passive stiffness is constant at more acute angles where the biceps brachii is further shortened. As the joint is ranged from flexion to extension, triceps brachii muscle stiffness is presumed to decrease until reaching its slack angle, then plateau. Concurrently, the biceps brachii is initially plateaued until reaching its slack angle where stiffness then increases. This superimposition of effects from antagonistic muscles likely resulted in the observed parabolic joint impedance profile as a function of elbow angle. The decreasing portion of the parabolic profile (90° to ∼110°), where elbow flexor stiffness is presumed constant (35), is likely due to decreasing stiffness of the elbow extensors up to their slack angle. Considering this assumption, one would expect that measurement of shear wave velocity in the long head of triceps brachii would be correlated with the decreasing portion of the joint impedance profile. Future work, with improved ability to measure shear wave velocity in the triceps brachii, is needed to confirm this postulation. Another explanation of the differing relationship of shear wave velocity with joint angular position is to consider the relationship between shear wave velocity and stress. Recent work has demonstrated that shear wave velocity provides a reliable estimate of stress in tendon (Martin J et al. 2018, Nature Communications, 1:10) and that shear wave velocity in passively stretched muscles depends largely on muscle stress (Bernabei et al. ‘Muscle stress provides a lower bound n the magnitude of shear wave velocity. International Society of Biomechanics Congress, Calgary, July 31-Aug 4, Calgary). Thus, the relationship between stress and joint position likely influences the shape of the shear wave velocity and joint angular position curve.

While shear modulus increased as a function of increasing elbow joint extension, it did not demonstrate the striking parabolic increase previously reported in normal biceps and brachialis muscle at end range (29). This may be explained by the present study not ranging the elbow joint through full extension. Therefore, the biceps muscle was not fully lengthened reaching the end of its elastic profile as defined by the Huxley model (27, 31). However, our range in passive muscle shear modulus was congruent with that reported by Gennisson et al of ∼5-15kPa within the range of 90° to 140°. In the present study, limiting extension to 150° was an unavoidable constraint that prevented undesired elicitation of hyperactive stretch reflexes, or spasticity, that is sensitive to both muscle length and velocity (36). The 150° end range position was determined empirically as the maximum elbow extension displacement that could be achieved across participants while preserving EMG quiescence. In addition, the biceps was not fully shortened beyond its slack angle where the shear modulus, or stiffness, would be expected to plateau. Measurement at angles more acute than 90° was not feasible in the present experimental setup since we were limited by the proximity of the arm to the torso within the experimental device and the needed space for the ultrasound transducer.

When comparing the limbs of individuals with stroke, the present study was consistent with previous reports of increased biceps stiffness (18, 39, 74) and elbow extension joint impedance (18, 43) in the paretic limb. However, one must exercise caution in this interpretation and acknowledge the primary limitation surrounding the comparison of arms. While exquisite attention was devoted to the confirmation of muscle quiescence, it is possible that undetectable flexor activation was occurring, especially toward end range of motion due to hyperactive stretch reflexes and/or hypertonicity. If so, this would have been reflected in all metrics and enhanced on the paretic side, additionally reflecting active as opposed to solely passive/intrinsic joint impedance. Other methods including ischemia (47), phenol motor point block (52), brachial plexus blockade (58), or sleep atonia (6) may fully suppress muscle activation and resolve this limitation in future research. Under the assumption of muscle quiescence, the present study found a 18% and 48% overall mean increase in shear wave velocity and shear modulus for the paretic arm, respectively. In comparison, Lee *et al* (39) reported a maximum of 69% increase in shear wave velocity and Wu *et al* (74) reported a 12-19% increase in the paretic biceps. The discrepancy in shear wave velocities may represent differences in sample populations between the studies. In the Lee et al. study, 4 of 16 reported participants had 2-to 3-fold increases in shear wave velocity on the paretic side. The remaining participants in the Lee et al. study reflected differences subjectively similar to the sample reported in the present study. In Wu *et al*, a cohort of 9 of 31 individuals with stroke were identified as having negligible differences between arms, comparable to controls yet reflecting values obtained in the present study. The range of differences across studies reflects the natural heterogeneity of stroke presentation. An alternative explanation for a discrepancy across studies would be the implementation of a standardized reflex habituation procedure employed in the present study. As discussed in the methods, 20 fast (120°/s) elbow extension-flexion stretches were conducted to habituate unwanted biceps stretch-reflex activation (68). This procedure was included in leu of the more laborious or invasive methods listed above since undetected muscle activation stands to have a substantial impact on shear wave velocity/shear modulus. In fact, shear modulus is highly sensitive to muscle activation and capable of differentiating activations of even 3% and 7% maximum voluntary activation (60), which is why the 3% threshold is indicated in Figure 4. To minimize undetected muscle activation as a source of error to the internal validity (71) of the experiment, the present study went beyond conventional visual EMG inspection by employing a quantitative threshold-based confirmation during post processing to confirm quiescence. While these rigorous steps bolster the validity of our methodology, they do not guarantee muscle quiescence, as can be achieved through methods of ischemia, motor point block, and sleep atonia. Therefore, studies of muscle stiffness and joint impedance implementing these techniques are needed.

The increase in shear wave velocity, shear modulus, and joint impedance on the hemiparetic side may be the result of longitudinal decreases in fascicle length (42) and/or changes in the expression of titin (48) and collagen (53). In the event of decreased muscle fascicle length in the hemiparetic biceps, the passive portion of the length-tension relationship would be shifted leftward similar to findings in the ankle by Gao *et al* (28), such that for a given joint angle, a larger increase in muscle tension would result. The methods employed in the present study use standardized joint angular position for measurement. Therefore, it is possible that the paretic biceps was positioned farther along its operational curve in comparison to the non-paretic side. Future investigations would benefit from standardizing by muscle fascicle length, perhaps with the assistance of extended view ultrasonography (59), to more accurately capture muscle and joint changes. This possible influence of fascicle length and also discrepancy in overall muscle length between the muscle of the paretic and non-paretic side may be reflected in recent findings of the strong relationship between shear wave velocity and stress in tendon (Martin J et al. 2018, Nature Communications, 1:10) and passive muscle (Bernabei et al. ‘Muscle stress provides a lower bound n the magnitude of shear wave velocity. International Society of Biomechanics Congress, Calgary, July 31-Aug 4, Calgary). Thus, the relationship between shear wave velocity and stress may contribute to differences observed between the paretic and non-paretic sides.

A compounding mechanism that may also be contributing to increases in shear wave velocity, shear modulus, and joint impedance in the paretic limb is a change in the amount or type of titin expressed within the myofibril and/or increases in the deposition of collagen in the extracellular matrix. While the exact roles of titin and collagen in passive muscle tension is still debated (44), it is widely excepted that the parallel elastic component contributes to the passive tension within the Hill-type model. Recent evidence reporting echogenicity in B-mode imaging supports alterations in muscle composition following stroke (39). Although it is unclear whether fascicle length and/or connective tissue changes explain the differences between limbs following stroke, it does not detract from the study objective of evaluating the *criterion* validity of ultrasound elastography in the paretic biceps that was further supported by all metrics detecting a significant effect of limb.

Interestingly, the measurement of joint impedance appeared to detect a greater impact of paresis than did shear wave ultrasound elastography. This was observed in a larger relative change in joint impedance compared to shear wave velocity and shear modulus between limbs. Joint impedance was 123% greater on the paretic side than on the non-paretic side whereas shear wave velocity was 18% greater on the paretic side. This difference may represent the summative effects of all muscles/tissues crossing the elbow joint that contribute to the joint impedance measure. Shear wave velocity was only measured in the biceps. The other elbow flexors making up a significant proportion of the cross-sectional area may undergo similar changes as well. An additional aspect may have been that measurement of joint torque did not account for the decreasing moment arm length in the biceps brachii that occurs with elbow joint extension beyond ∼90° (3, 5). A decreasing moment arm during elbow extension would serve to attenuate the contribution of increasing passive muscle force due to lengthening on passive joint torque. The degree to which the changing moment arm and passive muscle force contribute to passive joint torque is likely to be participant-specific and require musculoskeletal modeling employing additional measured parameters due to structural, functional, and metabolic changes that occur in muscle following stroke (67). While this was beyond the scope of the present study, future work may include measuring passive joint impedance by employing imaging to quantify in vivo participant-specific moment arms, muscle cross sectional area, and pennation angles as a function of joint angular position (2, 3, 37, 57).

More quantitative tools to evaluate changes in passive muscle mechanical properties are necessary for the advancement of current clinical methods. Clinical assessment of passive muscle properties are limited to palpation, joint mobility, and muscle length testing (63), along with resistance to lengthening due to abnormal stretch reflex, or spasticity (9). Importantly, clinical assessment of spasticity cannot differentiate the two constituents of total passive stiffness; that is, reflex stiffness and intrinsic stiffness (54). One commercially available instrument, *Myotonometer* (Neurogenic Technologies, Inc, USA), is capable of measuring tissue deformation and therefore muscle stiffness (15, 40, 41) with established reliability (15); however, it is limited in application in individuals with more adipose tissue. In this respect, shear wave ultrasound elastography may be applicable to a greater proportion of the stroke population. In stroke rehabilitation, the ability to quantify muscle mechanical properties would provide clinicians with a powerful tool to evaluate contributors to movement dysfunction. Pathological increases in muscle stiffness will only compound abnormal neuromuscular performance impairments such as reduced reaching range of motion that occurs as a function of abduction loading (21, 23, 73). Quantifying changes in muscle through the implementation of shear wave ultrasound elastography will identify new therapeutic targets, precipitating the expansion of conventional approaches for ameliorating movement dysfunction in stroke recovery.

## Data Availability

Processed data are available upon request from the corresponding author following printed publication of the manuscript.

## Acknowledgements

We would like to acknowledge Ana Maria Acosta, PhD for her contributions to the signal analysis software. Additionally, we would like to acknowledge the participants with stroke whose continued dedication to clinical research is responsible for the successful completion of this study.

## Grants

This work was supported by a National Institute of Child Health and Human Development R01 Grant (HD 84009: PIs−Dewald/Murray, CoI−Ellis) and K25 Grant (HD096116; PI: Gurari).

## Disclosures

None of the authors have financial or other relationships that might lead to a perceived conflict of interest.

## Notes

### Competing Interest Statement

The authors have declared no competing interest.

